# Validation of a HIV Whole Genome Sequencing Method for HIV Drug Resistance Testing in an Australian Clinical Microbiology Laboratory

**DOI:** 10.1101/2023.07.05.23292232

**Authors:** Frances Jenkins, Thomas Le, Rima Farhat, Angie Pinto, M Azim Anzari, David Bonsall, Tanya Golubchik, Rory Bowden, Frederick J Lee, Sebastiaan Van Hal

## Abstract

Detection of HIV drug resistance is vital to successful anti-retroviral therapy (ART). HIV drug resistance (HIVDR) testing to determine drug resistance mutations (DRMs) is routinely performed in Australia to guide ART choice in either newly diagnosed people living with HIV or in cases of treatment failure. In 2022, our Australian clinical microbiology laboratory sought to validate a Next-generation sequencing (NGS)-based HIVDR assay to replace the previous Sanger sequencing (SS)-based ViroSeq assay. NGS solutions for HIVDR offer higher throughput, lower costs and higher sensitivity for variant detection. We sought to validate the previously described low-cost probe-based NGS method (veSEQ-HIV) for HIV-1 recovery and HIVDR testing in a diagnostic setting. The implemented veSEQ-HIV assay displayed 100% and 98% accuracy in major and minor mutation detection respectively and 100% accuracy of subtyping (provided >1000 mapped reads were obtained). Pairwise comparison exhibited low inter-and intra-run variability across the whole genome (Jaccard similarity coefficient [J] =0.993; J=0.972) and limited to the Pol gene only (J=0.999; J=0.999) respectively. The veSEQ-HIV assay met all our pre-set criteria based on the WHO “Recommended methods for validating an in-house genotyping assay for surveillance of HIV drug resistance” and has successfully replaced the ViroSeq assay in our laboratory.Scaling-down the veSEQ-HIV assay to a limited batch size and sequencing on the Illumina iSeq100, allowed easy implementation of the assay into the workflow of a small sequencing laboratory with minimal staff and equipment and the ability to meet clinically relevant test turn-around times.

## Introduction

HIV is a major public health concern worldwide. While Australia has a low prevalence of HIV in the general population (0.1% on 2017 estimate), HIV detection and management still play an important role in transmission prevention especially in at-risk populations (Guy, 2007; Kirby Institute, 2018). HIV drug resistance (HIVDR) testing to determine drug resistance mutations (DRMs) is routinely performed in Australia to improve chances of treatment success by informing anti-retroviral therapy (ART) choice in newly-diagnosed people living with HIV or in cases of treatment failure.

Royal Prince Alfred Hospital in Sydney, Australia routinely performs HIVDR testing and from 2016 until 2021, utilised the Sanger sequencing (SS) based commercial assay, ViroSeq HIV-1 genotyping system (Celera Diagnostic, Abbott Laboratories, Alameda, CA, USA). The ViroSeq system includes sample purification and reverse - transcription polymerase chain reaction steps, to generate a 1.8 kb amplicon covering parts of the polymerase (*Pol*) region including the protease (PR) and partial reverse transcriptase (RT) genes. The generated amplicons act as the SS template to generate an approximate 1.2kb chromatogram which after manual curation acts as the sequence input to the HIVdb Program available on the Stanford University HIVDR database website (https://hivdb.stanford.edu/hivdb/by-patterns/). The HIVdb Program identifies known DRMs and inferred levels of drug resistance to commonly used anti-retroviral drugs (Liu TF, 2006; Rhee, 2003; Shafer, 2006).

In 2020, our laboratory was advised that the ViroSeq system would cease manufacture in mid-2021. We therefore sought to replace this assay with a suitable alternative that would be able to detect DRMs in samples with a plasma viral load around 2000 copies per mL (cp/mL) at similar costs and overcome several limitations of the ViroSeq system: (1) able to obtain results across all HIV-1 subtypes; (2) include the integrase (IN) region; 3) able to detect subconsensus variants as several studies have demonstrated that these subpopulations contribute to negative clinical outcomes (Metzner, 2009; Simen, 2009); and 4) was future-proof as novel ART agents become available. Next-generation sequencing (NGS) solutions for HIVDR have been proposed as superior to SS methods due to higher throughput, lower costs and improved sensitivity to detection minority variants at lower levels (Lee, 2020; Taylor, 2019). Several NGS-based HIVDR methods have been reported, however, the challenge remains finding protocols that work consistently without sampling bias across all HIV-1 subtypes and viral loads.

A robust low-cost probe-based high-throughput NGS method for HIVDR and viral load (veSEQ-HIV) was previously described by Bonsall et. al (2020) (Bonsall, 2020). Previous evaluation of its performance in comparison to ViroSeq, showed a high degree of concordance in DRMs detected, comparatively higher sensitivity in detection of sub-consensus variants and the ability to detect complete sequences in a majority of samples (70%), increasing with viral load (Fogel, 2020).

As such, the veSEQ-HIV assay best met our requirements. However, the veSEQ-HIV assay was designed to run in excess of 96 samples per batch (Bonsall, 2020). In our diagnostic setting, with a need to meet clinically relevant turn-around times, we reduced this batch size to 8 patient samples and performed sequencing on the Illumina iSeq100. These adjustments allowed us to implement the assay into the workflow of our small sequencing laboratory with minimal staff and equipment. Here, we describe the validation of the veSEQ-HIV method in its utility for HIVDR testing as an in-house IVD in an Australian clinical microbiology laboratory.

## Materials and methods

### Ethics

Human Research Ethics Committee of the Sydney Local Health District approved the use of stored plasma samples in this study (2019/ETH01706).

#### Study population/Sample selection

Sixty-six stored plasma samples with known DRMs from both treatment naïve and - experienced people living with HIV-1 that had undergone testing on the ViroSeq system between 2016-2021 were included. Associated plasma viral loads were obtained from laboratory records and ranged from 950 to >10^6^ cp/mL. Subtypes included B (n=48), C (n=3), CRF01_AE (n=13) and CRF02_AG (n=2). Isolate baseline characteristics are summarized in **Table 1**.

**Table 1.** Isolate baseline characteristics

| Subtype | Number of samples | Mean viral load (cp/mL) | Viral load range | Number Surveillance Drug Resistance Mutations |  |  |
| --- | --- | --- | --- | --- | --- | --- |
|  |  |  |  | PR** | RT** | IN |
| B | 48 | 364275 | 2260-2914185 | 6 | 54 | 5* |
| C | 3 | 41444 | 14309-81222 | 0 | 5 | 0* |
| CRF01_AE | 13 | 167506 | 1700-847000 | 0 | 26 | 0* |
| CRF02_AG | 2 | 53025 | 1050- 105000 | 0 | 4 | 0* |

\* Only 5/66 (8%) samples had integrase testing at baseline. Integrase is not included in ViroSeq assay.
\*\* Surveillance Drug Resistance Mutations as determined by ViroSeq assay.

As the ViroSeq assay does not cover the IN region, we included five samples that had had SS-based IN testing performed at an external accredited laboratory and had Stanford HIVdb reports available. We did not have access to nucleotide sequence for these samples and only included them in our comparisons of subtype and mutation detection.

For reference, a dilution series (10^5^ – 25 cp/mL) of the highest AcroMetrix HIV-1 Panel standard was prepared (Thermo Fisher Scientific, Waltham, Massachusetts, USA). A negative control (PCR confirmed HIV-negative plasma) was taken though the entire process from extraction to sequencing for each run.

### Nucleic acid extraction

Methods were based on those described for veSEQ-HIV (Bonsall, 2020) with minor modifications. Briefly, automated total nucleic acid (NA) extraction was performed using the NucliSENS e-MAG (Biomerieux, Marcy-l’Étoile, France). Extraction was performed using the manufacturers general extraction protocol with an input volume of 500 µL and elution of 25 µL.

### cDNA generation and library preparation

NA was concentrated using RNAClean XP beads (Beckman Coulter, Brea, California, USA) before quarter reactions for cDNA generation without RNA fragmentation were prepared using the SMARTer Stranded Total RNA-Seq Kit v2 – Pico Input Mammalian Kit (Takara Biosciences USA, San Jose, California, USA) with maximal NA input.

Libraries were prepared using the same kit. Indexed libraries were pooled equally by volume (8 patient and 1 negative control) before performing clean-up using a ratio of 0.68 Ampure XP beads (Beckman Coulter, Brea, California, USA). DNA quantification was performed using Qubit HS DNA kit and Qubit fluorometer (Thermo Fisher Scientific, Waltham, Massachusetts, USA) and fragment size determined using the HS D1000 assay on Tape Station 4200 (Agilent Technologies, Santa Clara, California, USA).

### Capture-based hybridization, post-capture PCR and clean-up

Up to 500 ng of pooled library was taken through viral capture using the xGen hybridization and wash kit (Integrated DNA Technologies, Singapore). The probe pool contained 646 custom 120-mer biotinylated probes targeting the HIV genome across multiple subtypes. The custom probe sequences are published here for the first time with permission (**Supplementary file 2**). Post-capture PCR was performed as per manufacturer’s instructions.

Clean-up was performed with a ratio of 1.5x Ampure XP beads (Beckman Coulter, Brea, California, USA). DNA quantification was performed using Qubit HS DNA kit and fluorometer (Thermo Fisher Scientific, Waltham, Massachusetts, USA) and fragment size determined using HS D1000 assay on Tape Station 4200 (Agilent Technologies, Santa Clara, California, USA). An additional post-capture PCR (6-cycles; total 16 cycles) was added and clean-up repeated if minimum library molarity (1nM for iSeq100) was not meet at the final library stage.

### Sequencing

Final libraries were prepared for sequencing as per the Illumina iSeq 100 Sequencing System Product Documentation (# 200015511 v00). A final loading concentration of 75 pM was used with a 2% PhiX control spike. Libraries were run on the Illumina iSeq100 system using iSeq 100 i1 Reagent v2 (300-cycles) with read lengths of 2 x 150 bp.

### Bioinformatics

Raw short-read data was processed by an in-house automated pipeline utilising open-source software as described previously with minor modifications (Bonsall, 2020). Briefly, Kraken was first used to filter out human reads followed by trimming of adapters using fastp. Host-depleted, filtered reads were then taken forward including generating assemblies using and SPAdes which formed the inputs to assemble the contigs suitable as input for SHIVER (v3) (Wood DE, 2019; Wymant, 2018).

An updated Python 3 compliant SHIVER was used for analysis setting a previously validated minimum depth of 5-15 against the reference genome (Human immunodeficiency virus type 1 - HBX2) (Ji, 2018). The resulting output is then run the Stanford University HIV Drug Resistance database to identify drug resistance mutations using the tool sierrapy (Liu TF, 2006; Shafer, 2006; Rhee, 2003). Subsequent reports are generated using an in-house script based on the Stanford json file output.

### Assessment of assay performance

Validation criteria were pre-defined and based on WHO “Recommended methods for validating an in-house genotyping assay for surveillance of HIV drug resistance” with minor modifications (WHO, 2020). In brief, these criteria included that 100% of known mutations must be detected and ≥90% of pairwise comparisons must be at least 98% identical. The WHO guidance also recommended meeting sensitivity for amplification ≥95% of samples with viral loads between 500 and 1000 cp/mL amplified and successfully genotyped (n ≥ 10). This criteria had to be modified as the ViroSeq (our “gold-standard”) is not suitable for viral loads <2000 cp/mL to sensitivity for amplification ≥95% of samples with viral loads >2000 cp/ml must be amplified and successfully genotyped (n ≥10). Performance metrics used to assess the new method were as follows: (1) accuracy; (2) precision/reproducibility; (3) sensitivity.

### Accuracy

Using sensitivity or specificity calculations in the context of sequencing comparisons are impractical and misleading as there are no “real” true negatives especially when including minor mutations in an RNA virus that has a low barrier for genomic change. Therefore, the Jaccard similarity coefficient (J) was employed which gauges the diversity and similarity of samples. The Jaccard coefficient measures similarity between finite sample sets, and is defined as the size of the intersection divided by the size of the union of the sample sets.

J(ViroSeq,WGS)=(| ViroSeq ∩WGS |)/(| ViroSeq∪WGS |)

By design values are between 0 and 1 with higher values indicating greater similarity.

The level of agreement between the new test and the reference test was assessed by comparing consensus nucleotide sequences, amino acid sequences, subtype, major and minor mutations detected. To compare nucleotide sequences, both WGS data and the Viroseq FASTA files were mapped to HIV-1 reference HBX2 using clustalW (Chenna, 2003). Relevant regions of the *Pol* region (1.3kbp) were extracted from the aligned files. Bases common to both datasets including ambiguous assigned bases in Viroseq data were extracted as well as mutations through pairwise comparisons.

### Linearity, Limit of Detection and Limit of Quantification

Both the limit of detection (LoD) and limit of quantification (LoQ) were assessed using a panel of standards comparing viral load to the number of reads mapping to the reference (HIV-1-HBX2). Although, the veSEQ-HIV assay is capable of estimating viral load based on reads mapping to the reference provided a panel of quantification standards is included in each run (Bonsall, 2020), this was not formally assessed given the predicted number of samples in our setting.

### Precision/Reproducibility

Inter- and intra-run agreement was assessed by running replicates on different runs and within the same run. For intra-run comparison, a set of five samples were included on the same run across three different runs; viral loads ranged between 3475 and 100,000 cp/mL and included subtypes B, C and CRF01_AE. For inter-run comparison, a set of eight samples were included on two different runs; viral loads ranged between 3435 and 100, 000 cp/mL and included subtypes B and CRF01_AE. A summary of the replicates can be found in **Table 2** below.

**Table 2.**
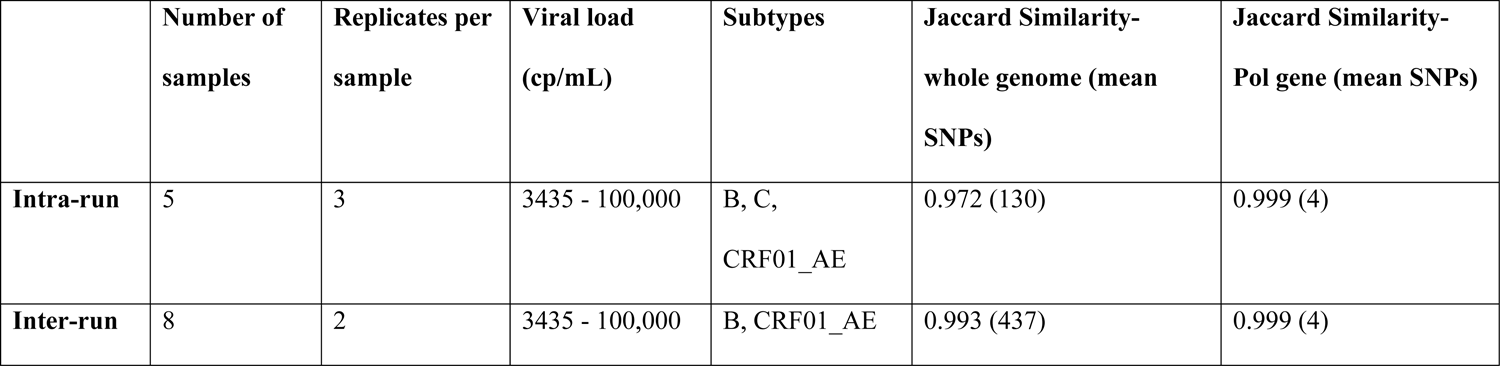
Intra- and inter-run comparison including Jaccard Similarity and SNP numbers across whole-genome and across *Pol* region

|  | **Number of**<br>**samples** | **Replicates per**<br>**sample** | **Viral load**<br>**(cp/mL)** | **Subtypes** | **Jaccard Similarity-**<br>**whole genome (mean**<br>**SNPs)** | **Jaccard Similarity-**<br>**Pol gene (mean SNPs)** |
| --- | --- | --- | --- | --- | --- | --- |
| **Intra-run** | 5 | 3 | 3435 - 100,000 | B, C,<br>CRF01_AE | 0.972 (130) | 0.999 (4) |
| **Inter-run** | 8 | 2 | 3435 - 100,000 | B, CRF01_AE | 0.993 (437) | 0.999 (4) |

The WHO recommends (3 x 5 replicates) to determine inter-run precision, due to limited remaining plasma from previously tested samples, inter-run precision was determined using a (2 x 8 replicates) approach.

### Specificity

Specificity was assessed by a negative control (PCR HIV negative plasma) included in all runs. Subsequent routine clinical runs have included a phosphate-buffered saline negative control. Possible interfering substances were not assessed as sequencing would only be performed on confirmed HIV positive samples.

### Quality control Criteria

A set of quality control (QC) criteria were established to assess both run and sample data quality prior to analysis. We included sequencing run metrics as specified by the manufacturer (Illumina) as well as criteria to review individual raw read data, inclusive of the negative control. For review of individual sample data, we referred to guidelines established by the Winnipeg Consensus for read quality control (QC), read alignment, reference mapping, HIVDR interpretation and reporting (Ji, 2018).

### Clinical Reporting

Development of our clinical reporting strategy involved discussion with treating clinicians in our network and well as incorporating requirements set by our accreditation, WHO recommendations and guidelines established by the Winnipeg Consensus (Ji, 2018; WHO, 2020).

## Results Accuracy

### Accuracy of nucleotide sequences

The pairwise analysis of the *Pol* region resulted in an overall Jaccard coefficient of similarity of 0.998, with 98,459 bases shared and 145 SNPs (single nucleotide polymorphisms) differences between isolate pairs.

### Accuracy of subtyping

Concordant subtypes were achieved for 98% (n=65/66) of patient samples. One sample was misidentified as subtype CRF_14_BG rather than subtype B. This sample had low viral load (2790 cp/ mL), resulting in less than 1000 mapped reads and suboptimal coverage across the PR gene. Provided samples obtained >1000 mapped reads 100% accuracy of subtyping could be obtained.

### Accuracy of Major Drug Mutations

Due to the low prevalence and success of current ART, a limited repertoire of major drug mutations could be sourced and included 5 different PR, 19 NNRTI/NRTI and 5 IN major mutations. There were 19 discordant major drug mutation calls between the two methods. All discordances were able to be resolved. The majority (n=9) on the ViroSeq chromatogram were assigned as an ambiguous base with reporting calling both wild-type and a DRM. These were all present on WGS but at a sub-consensus level (defined when <50% but >5% of reads mapping to the alternative allele at a >20 fold read depth). The remaining discordance were as a result of: (1) incorrect manual base assignments on ViroSeq (n=5); and (2) no coverage across region of interest on WGS (n=5). No false positive DRMs were detected on WGS. Taking these additional results into account, the overall accuracy/concordance for detection of major mutations was 100% with a Jaccard similarity coefficient of 1.0. A summary of these results can be found in **Table 3**.

**Table 3.** Concordance of major drug mutations between methods

| **Major mutation location** | **Concordant mutation calls** | **Discordant mutation calls** | **Presence of sub-population** | **ViroSeq poor coverage/false calls** | **WGS poor coverage** | **Discordance resolved (%)** | **Overall accuracy** |
| --- | --- | --- | --- | --- | --- | --- | --- |
| **PR** | 6 | 3 | 1 | 2 | - | 100<br>(n=3/3) | 100%<br>(n=6/6) |
| **RT** | 84 | 16 | 8 | 3 | 5 | 100<br>(n=16/16) | 100<br>(n=84/84) |
| **IN** | 5* | - | - | - | - | - | 100<br>(n=5/5) |
| **Total** | - | - | 9 | 5 | 5 | 100<br>(n=19/19) | 100<br>(n=95/95) |

Please note: Replicates were included in mutation analysis
\* SS IN results only available for 5/66 samples

### Accuracy of Minor Drug Mutations

Only accuracy for PR and RT were examined. Three (3/601; 0.4%) and 29 (29/1249; 2%) discordant minor PR and RT mutation calls were identified respectively while 1 (1/569; 0.2%) and 13 (13/2499; 1%) potential WGS false positives were identified. The majority of the false positive (n=10) minor mutations occurred on the boundaries of the ViroSeq amplicons resulting in higher uncertainty of basecalls. Irrespective, the overall accuracy/concordance for detection of Minor mutation was 98%. The overall Jaccard coefficient of similarity was 0.975. A summary of concordance of minor drug mutations is available in **Supplementary Table 4**.

### Limit of Detection and Limit of Quantification Mapped reads

Eight samples (8/82; 10%) run on WGS had suboptimal coverage (<1000 mapped reads). Five (5/8; 63%) of these samples had low viral loads (2000 - 5000 cp/mL). The depth of coverage affected all regions when <1000 mapped reads were obtained while >1000 mapped reads gave excellent coverage depth for all regions. Results are summarised in **Table 4**.

**Table 4.** WGS QC metrics based on number of mapped reads

| No. mapped Reads | No. of samples | Average Depth Coverage |  |  |
| --- | --- | --- | --- | --- |
|  |  | PR region | RT region | IN region |
| <1000 | 8 | 71% @ 9 fold | 42% @ 8 fold | 52% @ 7 fold |
| >1000 | 78 | 99% @ 2914 fold | 99% @ 3465 fold | 99% @ 3372 fold |

### By viral load

A significant linear correlation (R=0.74; p<0.01) between viral load and number of mapped reads. (Figure 1A and 1B) was detected. Low viral loads (2000 - 5000 cp/mL) gave inconsistent results but were linked to the number of mapped reads with <1000 mapped reads in six of 13 isolates (46%). The remaining seven represented possible failed WGS runs.

**Figure 1A& 1B.**
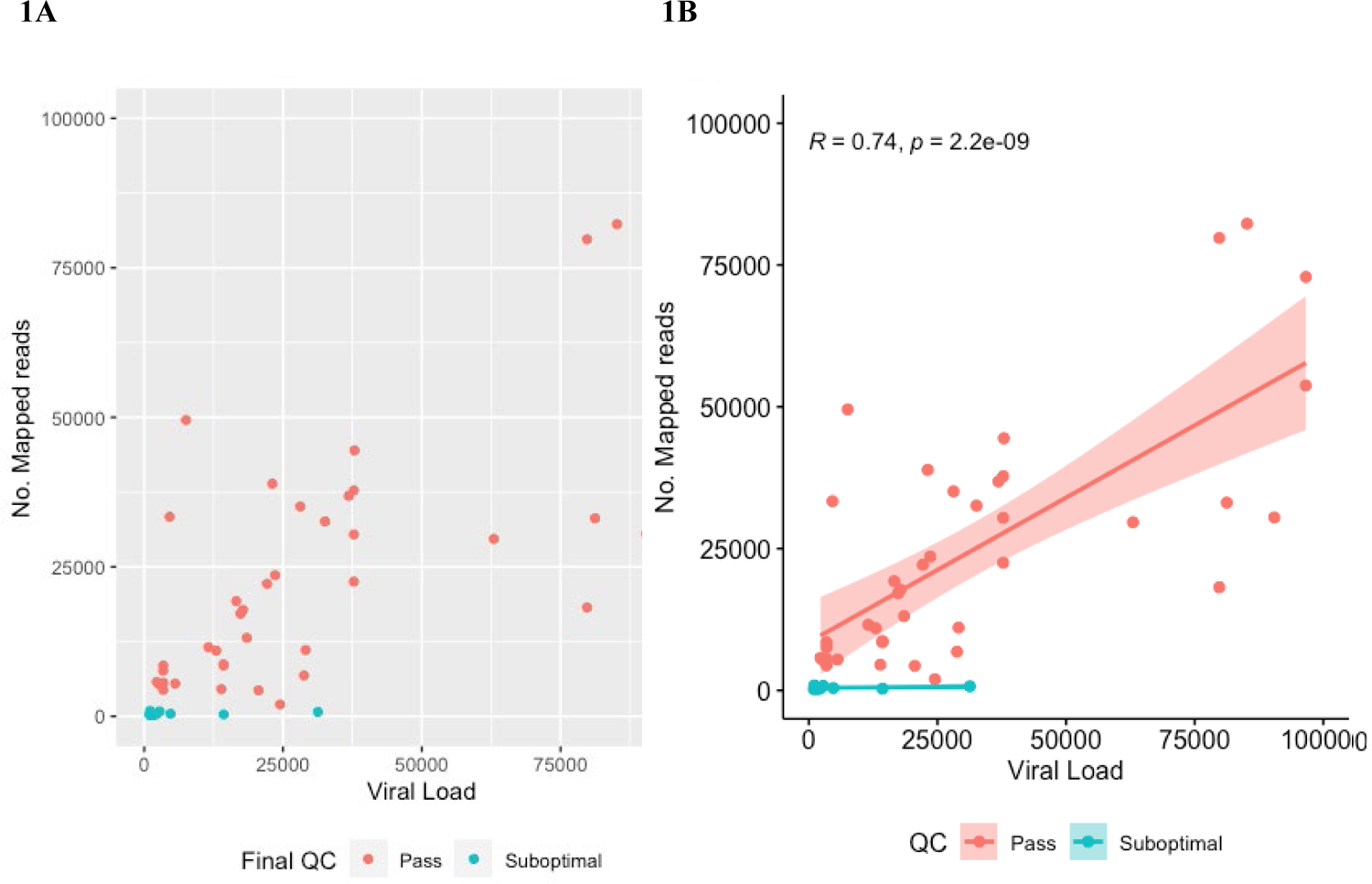
Scatterplot of viral load and number of mapped reads for sequenced isolates (1A) and correlation and prediction curve (1B).

These were unable to be repeated as no further sample was available. Furthermore, as all samples had been stored with an unknown number of freeze-thaw cycles, re-quantification was not possible to exclude sample RNA degradation as the cause.

### Linearity

A significant linear relationship (R=0.99; p<0.001) was detected when including a commercial standard AcroMetrix HIV-1 Panel (Thermo Fisher Scientific, Waltham, Massachusetts, USA). With decreasing viral loads a reduction in: (1) the percentage coverage across genes; and (2) the number of mutations was detected. At low viral loads <2000 cp/mL potential false positive (FP) mutations were detected (**Supplementary Table 1**). A limit of detection for HIV-1 was determined to be 100 cp/mL. With respect to reporting the number of mapped reads was the best correlate with optimal reporting requiring a minimum of 1000 mapped reads.

### Inter-run (reproducibility) and intra-run (repeatability) of WGS method

The inter-run variability (good Jaccard similarity index) both across the whole genome (J=0.993) as well as across the *Pol* gene (J=0.999) was within the expected performance with an average of 437 SNPs across the whole genome and 4 SNPs across the *Pol* gene between runs.

Similarly, the intra-run variability (good Jaccard similarity index) was adequate both across the whole genome (J=0.972) as well as across the *Pol* gene (J=0.999) with an average of 130 SNPs across the whole genome and 4 SNPs across the *Pol* gene between runs. These results are summarised in **Table 2**.

### Specificity

No evidence of contamination across all validation runs or subsequent clinical runs was detected with an average of 20 (range 0-548) reads for the negative control. Although reads represented HIV reads in a small subset, all negative control assemblies failed and mapping to the reference (HIV-1-HXB-2) was unsuccessful with 0% coverage and 0% depth across the three regions (PR, RT, IN) of the HIV genome.

### Quality Control Criteria

QC metrics and their corresponding limits/criteria for sequencing runs are presented in

## Supplementary Table 2

### Clinical reporting

In the transition to NGS-based methods, WHO has recommended presenting resistance results in the same way as previous SS-based methods (WHO, 2020). Thus, reports include the same data output generated by the ViroSeq assay-subtype identification and lists identified DRMs mutations and their associated susceptibility interpretations.

With interest expressed from the treating clinicians in our network, the presence of sub-consensus variants containing major drug resistance mutations are also included (reporting when frequency greater than 5% and with appropriate caveats) (Ji, 2018). These findings are presented in an iterative way with additional mutation score and interpretation change reported alongside consensus interpretations. With the clinical relevance of sub-consensus variants of <20% frequency still under debate, this leaves the choice to consider any changes in interpretation to the treating clinician. An example report can be found in **Supplementary file 1**.

## Discussion

In this study, we provide our in-house validation of the veSEQ-HIV assay in its utility for HIVDR in an Australian clinical microbiology laboratory. The veSEQ-HIV assay with minor modifications, met our pre-specified validation criteria based on the WHO “Recommended methods for validating an in-house genotyping assay for surveillance of HIV drug resistance” (WHO, 2020). We therefore deemed the assay fit for purpose and able to replace the ViroSeq assay in our laboratory.

We achieved a Jaccard similarity coefficient (J) of 0.998 for pairwise nucleotide comparisons of the *Pol* gene and detected 100% of major mutations (Criteria 1: Accuracy 100% of known mutations must be detected).

A limitation of our validation includes the small number of mutations encountered. We only encountered (11/56; 19%) NRTI; (8/49; 16%) NNRTI, 5/57; 9% PR and 3/43; 7% IN DRMs (All DRMs encountered are listed in **Supplementary Table 3**). Ideally, our validation set would have included all known mutations that affect clinical interpretation i.e. all known DRMs, however, practically, this is infeasible as hundreds of mutations would need to be covered (WHO, 2020). We attempted to cover as many mutations as possible but were limited to samples that had previously been referred to our laboratory for testing that had sufficient remaining plasma. Ongoing clinical testing should add to the DRMs encountered.

Pairwise comparison exhibited low inter-and intra-run variability across the whole genome (J=0.993; J=0.972 respectively). Agreement was highest when limited only to the *Pol* gene (both J=0.999). (Criteria 2: ≥90% of pairwise comparisons must be at least 98% identical). This supports previous finding of relative conservation across the *Pol* gene relative to the rest of the genome (Troyano-Hernáez, 2022). We identified 100% of subtypes correctly (Criteria 3: Sensitivity for amplification: ≥95% of samples with viral loads >2000 cp/mL must be amplified and successfully genotyped (n ≥10)).

There were 4 subtypes (B, C, CRF01_AE and CRF02_AG) represented in our sample set. The majority of samples were subtype B (n=48/66; 72%) followed by subtype CRF01_AE (n=20/66; 18%). This is in keeping with previous Australian epidemiological data identifying subtype B as the most prevalent and CRF01_AE as one of the dominant non-B subtypes of increasing prevalence (Castley, 2017). This indicates that while we could not equally represent all HIV genotypes due to lack of sample availability the subtypes in our sample are representative of circulating HIV in Australia. Irrespective, we do not expect there to be any negative bias in “uncommon” genotypes as veSEQ-HIV probes have been designed and previously shown to consistently recover genomes from these “uncommon” subtypes including C, A (A1 &A2), D, G and J (Bonsall, 2020).

The veSEQ-HIV assay is now in routine use in our laboratory and has been used to successfully sequence and generate over 100 HIVDR and genotyping reports to clinicians across the state of New South Wales, Australia.

Running limited batches (eight patient samples) has allowed easy implementation of the assay into the workflow of our small sequencing laboratory with minimal staff and equipment and has been an ideal use of a small desktop sequencer such as the Illumina iSeq100. Despite small batches, reagent cost per assay is still significantly reduced compared to the previous commercial assay (from $360 to $200). Assay batching flexibility also allows for future throughput increases utilising automated liquid-handing for library preparation and larger sequencing platforms (e.g. Illumina MiSeq).

Processing of NGS data has been identified as a major hurdle in implementation of NGS-based HIVDR. The Winnipeg Consensus has established preliminary guidelines for read quality control (QC), read alignment and reference mapping, variant calling and QC, HIVDR interpretation and reporting as well as general analysis of data management (Ji, 2018). These guidelines were invaluable in developing our bioinformatics, QC and clinical reporting strategy.

Further hurdles for NGS-based HIVDR include lack of specifically developed external quality assessment (EQA) strategies and programs (Lee, 2020; Ji, 2018) as current HIVDR EQA are still designed for SS-based methods. In the transition to NGS-based methods, WHO has recommended configuring NGS methods to present results in the same way as SS-based methods (WHO, 2020). Thus, while not ideal, NGS-based HIVDR can still be used to produce results formatted as SS-based HIVDR outputs, permitting participation in current EQA programs such as the HIVDR programs from Quality Control for Molecular Diagnostics (QCMD) that our laboratory participates in.

Informal sample exchange between laboratories offers an alternative transitional EQA strategy to meet accreditation needs, however, very few laboratories are currently performing NGS-based HIVDR in Australia. We look forward to more laboratories taking up NGS-based methods to make this another transitional EQA option.

Despite this, there remains an urgent need for appropriate EQA programs and materials to be made available as these approaches overlook the abundance and available complexity in NGS HIVDR data, particularly its ability to detect sub-consensus variants at low levels (Lee, 2020).

In addition, appropriate reference materials for validation of sub-consensus variant detection do not yet exist. Our reporting strategy for sub-consensus variants followed the recommendations from the Winipeg consensus with a 5% frequency threshold suggested to account for errors/bias introduced during assay steps such as reverse-transcription, PCR, sequencing steps etc. (Ji, 2018; Casadellà, 2017). While we did not quantify the sensitivity of variant detection in this validation, this threshold is likely conservative with estimates of minority variant frequency likely to be more robust using the probe-based veSEQ-HIV method as biases introduced by PCR are minimised and contamination is computationally controlled (Bonsall, 2020). Our reporting includes necessary caveats to sub-consensus variant reporting and leaves the choice act on subsequent drug susceptibility interpretation changes to the treating clinician.

Well-characterised sample panels including both clinical and synthetically constructed samples containing sub-consensus variants are needed to assess each laboratory’s ability to detect sub-consensus variants accurately (Lee, 2020).

## Conclusion

The veSEQ-HIV assay provides a robust and cost-effective NGS method for HIVDR in a diagnostic setting. veSEQ-HIV demonstrated satisfactory and consistent performance including low inter-and intra-run variability and good accuracy in detection of DRMs in samples with >1000 mapped reads (achievable in samples with VL >1000 cp/mL) when compared to the “gold-standard” SS-based ViroSeq assay. The veSEQ-HIV assay met all our pre-set criteria based on the WHO “Recommended methods for validating an in-house genotyping assay for surveillance of HIV drug resistance” and has successfully replaced the previous SS-based ViroSeq assay in our laboratory.

The assay meets current needs for clinical reporting (as SS-like results) to meet requirements of our diagnostic laboratory accreditation and to participate in current EQA programs as well as the ability to report on the presence of sub-consensus variants with appropriate caveats. As HIVDR transitions from SS-based methods to NGS and the clinical relevance of sub-consensus variants is further clarified, the veSEQ-HIV meets both current and future needs for HIVDR testing in a diagnostic setting.

## Data Availability

All data produced in the present study are available upon reasonable request to the authors

## Acknowledgements

Thanks to Alicia Beukers for her preliminary work in this validation. Thanks also go to the Clinical Immunology Laboratory at RPAH for providing the study samples and sharing the ViroSeq comparison data. Thanks to the Oxford group for providing the custom probe sequences to allow us to perform the veSEQ-HIV in our laboratory.

## Contributions

FJ drafted the manuscript.

FJ, TL, RF performed the validation testing

SVH, AP, FJL designed the validation

SVH developed the in-house pipeline, performed the bioinformatics and statistical analysis

MAA developed the custom probe sequences

TG assisted with pipeline development

DB, RB shared protocols and assisted with assay implementation

DB, RB, SVH, AP, FJL reviewed the manuscript

## Appendix. Supplementary materials

Supplementary file 1: Example Clinical Report

Supplementary file 2: Probe sequences

Supplementary file 3: Supplementary data

