## Supplementary File 3 for "Validation of a HIV Whole Genome Sequencing Method for HIV Drug Resistance Testing in an Australian Clinical Microbiology Laboratory"

### Supplementary Table 1. WGS results of dilution series of commercial standard AcroMetrix HIV-1 Panel

| Estimated VL | Mapped reads | Subtype | PR % | RT % | IN % | No. Mutations detected |
|---|---|---|---|---|---|---|
| 1000000 | 937160 | B | 99 | 100 | 100 | 22 |
| 100000 | 47356 | B | 99 | 100 | 100 | 22 |
| 10000 | 8766 | B | 99 | 100 | 100 | 22 |
| 1000 | 1086 | B | 99 | 83 | 65 | 19 & 1 FP |
| 100 | 114 | B | 46 | 18 | 32 | 1FP |
| 50 | 10 | B | 12 | 0 | 0 | 0 |
| 25 | 20 | B | 75 | 0 | 0 | 0 |
| 0 | 6 | CRF01_AE | 0 | 0 | 0 | 0 |

Supplementary Table 1 depicts the quantification and limit of detection against the commercial standard and sequenced using WGS. The Standard HIV sample (subtype B with drug susceptible profile) contained 20 minor/accessory mutations [PR = 3; RT = 13 and IN = 4].

**Supplementary Table 2. Selected QC metrics for sequencing run/sample analysis**

| **QC metric** | | |
|---|---|---|
| **Illumina sequencing run (iSeq100 i1 v2 reagent; 300 cycles;2 x 150bp)** | Occupancy (%)[*] | ≥80[**] |
| | Clusters passing filter (%PF)[*] | ≥53[**] |
| | Overall Qscore (%) | ≥80%^ |
| | PhiX error rate (%) | ≤1 |
| **Negative control** | Mapped reads (HIV-1-HXB-2) | <1000 |
| **Sample quality** | Mapped reads (HIV-1-HXB-2) | >1000 |
| | Average mapped read quality | >Q30 |
| **HIV-1 genes (PR, RT, IN)[#]** | Gene coverage (%) | >50 |
| | Read depth | >15 |

[*] On the Illumina iSeq100, %PF and % Occupancy represent an expected level to maximize data output and data quality. Metrics may fall outside of these ranges but still produce acceptable data as quality can still be high even with a lower quantity of reads. (Bruzek et al. 2020)

[**] QC metric determined by quantification of mean – 3SD for runs included in the validation

^ Manufacturer sequencing specifications

[#] The confidence of excluding HIV-1 gene resistance mutations (i.e. determining drug-class susceptibility) increases with overall gene coverage (>50%) and read depth (>15).

**Supplementary Table 3. Major and minor mutations encountered in our sample set**

| Region | Major mutations | Minor mutations |
|---|---|---|
| **PR** | M46L; G48A; I54V; V82A; L90M | K14R; T12S; N37I; L63P; I62V; T12P; I13V; R41K; E35D; I15V; L10I; T12R; V77I; G16A; P39S; G17E; T12A; T12K; G16E; T4S; T4P; L10V; L19I; T12N; N37S; R41K; I15V; I62V; Q92K; L63P; K45R; N37E; N37S; I13V; L19I; V11A; K20R; E35D; L10I; T12K; M36I; L19Q; L33V; L19T; G16E; L33I; T12A; R57K; K14R; I93L; V77I; Q18H; D60E; G17E; L63Q; L63T; G16A; L63H; Q61E; H69K; L19V; N37A; I64V; N37D; N37T; L63A; I72V; K20M; H69C; I64L; T74A; L63S; N37V; K20I; A71T; E65D; A71V; K43R; M36T; C67S; L89M; I72T; H69Y; N37K; M36V; T74S; H69Q; K70E; L63V; R41N; K70R; Q58E; V82I; L89I; I85V; T91A; I72M; T91V; F99L; I93M |
| **RT** | M41L; K65R; D67N; T69D; K70E; K70T; V75M; V75I; L100I; K103N; E138A; E138Q; Y181C; M184V; G190A; T215D; K219Q; P225H; N348I | K20R; K32E; D123E; V35I; S68G; E6D; A158S; V35T; I50V; V8I; R83K; E6A; V60I; V35L; K22R; I142L; V245R; K122E; K102R; K11R; S48E; P4Q; I135T; A98S; K11Q; D86E; V35T; K173Q; I135V; K103R; K122Q; S162A; T39L; V60I; K64R; K11I; D121H; S68G; K101Q; T39K; Q207E; A98S; K166R; K11T; K49R; D177E; I135T; T200A; S48T; I293V; K104R; R83K; K20R; E40D; T69N; T39S; E6D; D123E; T39R; F171Y; T69A; V35I; V8I; E399D; E291D; L533M; K390R; E404D; S468P; L469I; K366R; G335D; E432D; V435E; K281R; A554S; V435I; Q480H; F346Y; K558R; E514Q; H483Y; K512R; I329V; M357K; D324E; T477A; H483N; N447S; Q334L; T386I; V292I; S519N; R461K; I326V; L491S; R356K; K431S; K530R; L452I; R358K; L469F; A400T; E514D; L503M; A554N; L491P; L517T; A376S; T403M; G359S; V531I; L491A; Q524E; E546D; A554T; A371V; D471E; V559I; H483Q; A534S; T377M; K431T; Q464K; A376T; T470A; E529D; G196E; K277R; V245I; S162C; V189I; I135L; Q174K; K122E; P170T; E248N; V245Q; Q207D; F214L; R172K; D123S; E312A; Q278E; A288S; R211K; I178L; V245E; I178M; M357T; A272P; R211Q; T200I; K43E; I202V; S134G; E169D; I142T; E298A; I142V; Q174N; T286A; K201R; Q334H; Q207A; D121Y; Q207H; T200V; D460N; D121C; L283I; V245M; V466I; T286P; I329L; V179I; K311R; K102Q; K275R; S379C; D250E; E396D; G333E; G436E; V317A; P294S; L228Y; Q334R; E344K; Q334E; T240K; R307K; S322T; F440Y; R211S; K238R; A360T; K350R; I393V; H315Y; A272G; T39A; P294T; E297K; F346C; E297A; E344D; |

| | | |
|---|---|---|
| | | E203D; T200E; L301I; K476Q; R284K; S379G; E297V; K173S; V381I; E312T; Q334S; V435A; H483S; A502V; T377L; Q520K; V467I; E298D; T377R; T403V; E449D; V458I; T470N; R358G; N418S; L517I; I506L; H483L; G490E; R463K; A554K; E516D; V548I; L452Q; I434M; V518I; L517V; I556V; K527N; S468T; Q524K; E492K; K550R; S489A; H483C; S515L; K527Q; L310I; A502G; E492A; R448K; K512Q; K451R; G359T; V435M; S468C; T403R; I434L; K527R; T377V; L452V; K476R; T450S; T386V; K395R; E370D; A400V; I375V; E370A; S251I; L228R; D123N; K173T; I178V; V245N; K173I; I135R; S162Y; I135K; K249R; K385R; V245K; P294Q; V245T; I257L; K173A; K173R; V365I; Q207K; P345Q; D324G; T369A; L325I; S468N; D320E; V276I; P345T; A376V; S322P; Q207S; T296S; L301M; T377Q; T200L; I411V; A400S; K512T; V245A; Q334P; L484I; T470Q; V435T; V417I; T377N; E312N; T369V; Y342F; V435L; E36A; G196R; T39E; K122P; E204Q; K43A; V118I; I94L; K11A; S251T; K173E; I132L; G196V; K102R; D177G; E53D; T39M; E204D; I382V; Q509K; E312S; A508G; L452A; Q394R; E297R; Q174T; G335S; T286V; E248D; E370V; E328D; K465R; F389L; Q524H; Q197H; T386A; A304E; S251C; Q394L; A272S; T470K; A534T; M357R; Q547K; T403I; G335E |
| **IN*** | N51Y; T97A; Y143C;<br><br>N155H; N155K | Not examined |

14 * Sanger Sequencing Integrase results only available for n=5 samples

**Supplementary Table 4. Concordance of Minor Drug Mutations**

| Minor mutation location | ViroSeq Total mutation calls | ViroSeq Discordant mutation calls | WGS Total mutation calls | WGS Discordant mutation calls |
|---|---|---|---|---|
| **PR** | 601 | 3 (0.4%) | 569 | 1 (0.2%) |
| **NNRTI/NRTI** | 1249 | 29 (2%) | 2499* | 13 (1%) |

* 1317 of these mutations are post amino-acid position 320 in the RT gene; a region not amplified in the Viroseq assay
