## Supplementary File 1 for "Validation of a HIV Whole Genome Sequencing Method for HIV Drug Resistance Testing in an Australian Clinical Microbiology Laboratory"

**Supplementary file 1: Example Clinical Report**

### Royal Prince Alfred Hospital

Missenden Road, Camperdown, NSW 2050
Department of Microbiology

**HIV-1 Sequencing Report**

**Name of Patient:**

**MRN:** **Date of Birth:**

**Date of Collection:** **Referring Doctor:**

**Provided HIV Viral Load :**

**Our Reference / Accession:** **Date of Analysis:**

**Referring Laboratory:** **Referring Lab. Reference:**

**Sequencing Contact Staff:**

**Clinical Enquiries:**

**Methodology:**

Capture based whole genome sequencing was performed on this sample. HIV-1 RNA was extracted followed by a one-step RT-RNA PCR prior to sequencing using Illumina technology. Consensus genome was generated using Shiver v 3 specifying a minimum depth of 5-15 against the reference genome (Human immunodeficiency virus type 1 - HBX2)[1]. These thresholds have been validated against previous Viroseq results and recommended bioinformatic strategies[2]. Drug resistance mutation(s) were subsequently identified using the web-interface tool sierrapy against the Stanford University db [version(s): Sierra 3.4.3 (2023-04-26)]. Variant frequency for major drug-resistance mutations are determined with sub-consensus variants (at a frequency > 5%) indicated in the table. Potential drug susceptibility changes are based on Stanford major resistance mutations.
The effect(s) on drug resistance with the inclusion of sub-consensus mutations are shown in the table(s) under "score +" and "interpretation change" column(s). Minor variants are reported only if represented in the majority-population.

1. Wymant C. et al. Virus Evolution, Volume 4, Issue 1, January 2018, vey007. https://doi.org/10.1093/ve/vey007
2. Ji H. et al. Journal of the International AIDS Soceity, 2018, 21:e25193. https://doi.org/10.1002/jia2.25193

**Results:**

**No. reads mapped:** 841138
**HIV-1 subtype:** B (84%)
**QC metrics across HIV-1 genes**

| Gene | Coverage (%) | Av. Depth (No. reads) |
|---|---|---|
| **PR** | 98.99 | 16556 |
| **RT** | 99.82 | 14471 |
| **IN** | 99.65 | 25440 |

The confidence of excluding HIV-1 gene resistance mutations (i.e. determining drug-class susceptibility) increases with overall gene coverage (>50%) and read depth (>15). Subsequent interpretations are based on aligned and sequenced data. Repeat testing in the setting of low viral loads (i.e. mapped reads) is unlikely to provide greater confidence.

**Drug-resistance mutations:**

| Mutation | Gene | Stanford Type | Total No Reads | Ref Coverage | Mutation Coverage (%) |
|---|---|---|---|---|---|
| **K101P/Q/T** | RT | NNRTI | 5153 | 3907 (75%) | 1316 (25%)* |
| **E138K** | RT | NNRTI | 7174 | 4309 (60%) | 2910 (40%)* |

Coverage corresponds to the number of reads supporting the reference and variant or deletion.
A few studies have been able to demonstrate sub-consensus variants (<20% frequency) contribute to negative clinical outcomes, however, the overall clinical relevance of detection of these variants is unknown.

**Drug-susceptibility interpretations - Protease (PR)**

| class | name | score | interpretation |
|---|---|---|---|
| PI | atazanavir (ATV/r) | 0 | Susceptible |
| PI | darunavir (DRV/r) | 0 | Susceptible |
| PI | fosamprenavir (FPV/r) | 0 | Susceptible |
| PI | indinavir (IDV/r) | 0 | Susceptible |
| PI | lopinavir (LPV/r) | 0 | Susceptible |
| PI | nelfinavir (NFV) | 0 | Susceptible |
| PI | saquinavir (SQV/r) | 0 | Susceptible |
| PI | tipranavir (TPV/r) | 0 | Susceptible |

Major / Accessory (PR) Mutations: None detected
Other Mutations: L10V; T12P; K14R; I15V; M36I; N37D; L63P; I93L

**Please note:** Drug-susceptibility interpretations shown in this example report only include the Protease region. Reverse Transcription and Integrase regions are reported in a similar manner.
